# On the anti-correlation between COVID-19 infection rate and natural UV light in the UK

**DOI:** 10.1101/2020.11.28.20240242

**Authors:** Arnon Blum, Constantina Nicolaou, Ben Henghes, Ofer Lahav

## Abstract

While it is well established that the rate of COVID-19 infections can be suppressed by social distancing, environmental effects may also affect it. We consider the hypothesis that natural Ultra-Violet (UV) light is reducing COVID-19 infections by enhancing human immunity through increasing levels of Vitamin-D and Nitric Oxide or by suppressing the virus itself. We focus on the United Kingdom (UK), by examining daily COVID-19 infections (F) and UV Index (UVI) data from 23 March 2020 to 10 March 2021. We find an intriguing empirical anti-correlation between log_10_(F) and log_10_(UVI) with a correlation coefficient of −0.934 from 11 May 2020 (when the first UK lockdown ended) to 10 March 2021. The anti-correlation may reflect causation with other factors which are correlated with the UVI. We advocate that UVI should be added as a parameter in modelling the pattern of COVID-19 infections and deaths. We started quantifying such correlations in other countries and regions.

## 1 Introduction

Our hypothesis is that natural UV light suppresses the spread of COVID-19 virus in at least two ways: the effect on the virus itself, and on the human skin. We note that while natural UV may cause skin cancer, it generates vitamin-D which supports the immune system.

There are three types of solar UV radiation classified according to their wavelength. UVC is a short wavelength (100−280 nm) radiation and it is the most damaging to the human body. However, it is completely filtered by the atmosphere and does not reach the Earth’s surface. It is well known that UVC produced in the lab is used to inhibit viruses. UVB is a medium wavelength (280−315 nm) radiation and most of it is filtered by the atmosphere. UVA is a long wavelength (315−400 nm) radiation and accounts for about 95% of the UV radiation reaching the Earth’s surface. The UV Index (UVI) is a measure of the strength of sunburn-producing UV radiation at a particular place and time. Typical values of UVI in the UK range between 0 and 8.

The possible correlation between UV light and COVID-19 has been discussed in the literature, with contradictory conclusions. One study [1] found no association of COVID-19 transmission with UV radiation in Chinese cities. Others [2] found modest impact of UV light and other environmental effects on the reduction of COVID-19 transmission. Another study [3] pointed out that UV radiation will not be effective in places with high air pollution, where UV light turns into heat. On the other hand, other studies [4–6] have found that UV light is associated with decreased COVID-19 growth rate. Given the disagreement on the impact of UV light on COVID-19 transmission we take a fresh look at data for the UK. Our study is also strongly motivated by the second wave of COVID-19 in many Northern hemisphere countries during their winter.

## 2 Background on the impact of UV

Most of the respiratory viral infections have a seasonal pattern, that may be related to climate changes, humidity and UV irradiation from the sun, latitude, air pollution, height, and the human nature (genetic and epigenetic factors and behavioural characteristics). Enveloped viruses have a cold temperature preference (influenza A and B) [7]. A study examined the climate of 50 cities that were affected with the COVID-19 found that 8 cities had high morbidity and mortality rates. All 8 cities were located between latitudes 30°N and 50°N, with a temperature between 5°C to 11°C, and low humidity. Countries located below latitude 35°N had lower COVID-19 mortality rates. Countries located above 35°N have insufficient sunlight necessary for vitamin D activation. Vitamin D deficiency was found to correlate with hypertension, diabetes mellitus, and obesity, and associated with increased mortality rates [8–11].

Countries that suffered the highest mortality are known to have a high prevalence of vitamin D deficiency (Italy, Spain, UK, France). In Nordic countries, where sunlight is limited, vitamin D food fortification is mandatory and the mortality rate was lower in the recent pandemic [12]. Milan latitude is 45°N, and Naples is located at 40°N. Naples gets 58 more sunny days annually compared with Milan [13]. In Naples the death toll from COVID-19 was 403/million compared with 15,729/million in Milan (a >39 fold increase). A study in Scotland found that UVA exposure was inversely associated with myocardial infarction incidence, without any relation to temperature or UVB irradiation [14]. UVA penetrates to the epidermis, reaching blood vessels, keratinocytes and endothelial cells [15].

Sunlight activates Nitric Oxide (NO) in the skin. NO is a potent modulator of the cardiovascular system, reducing blood pressure and peripheral resistance [16–18]. NO is a signalling molecule responsible for the hoemostasis of blood vessels, affects cellular proliferation, inflammatory processes, and has an anti-bacterial and anti-viral activity [19–21]. Usually NO is produced in endothelial cells by oxidation of L-arginine to NO and citrulline. However, inducible NOS (i-NOS) is calcium-independent, activated during stress conditions like acute or chronic inflammation or infection [22–25]. NO inhibited the replication of SARS-CoV by inhibiting fusion between the S protein and its receptor, the angiotensin converting enzyme to receptor and through inhibition of viral RNA replication [26].

UVA penetrates into the dermal layer through the keratinocytes to the fibroblasts and the microvascular endothelial cells [27]. Keratinocytes, Langerhans cells, dermal fibroblasts, and melanocytes – all these cells have the ability to induce i-NOS once these cells were activated by cytokines [28].

## 3 Results

In our study we focus on the UK, using COVID-19 data [29], and UVI data for London [30]. Although the UVI is for London, the variations in different places in the UK are within 0.8 UVI. Stringency index represents the lockdown measures taken in the UK and defined in [31]. Figure 1 shows the daily infections and deaths for the UK along with the UVI and stringency index for the period 22 January 2020 to 10 March 2021. We visually observe a strong anti-correlation between daily infections and UVI.

**Figure 1:**
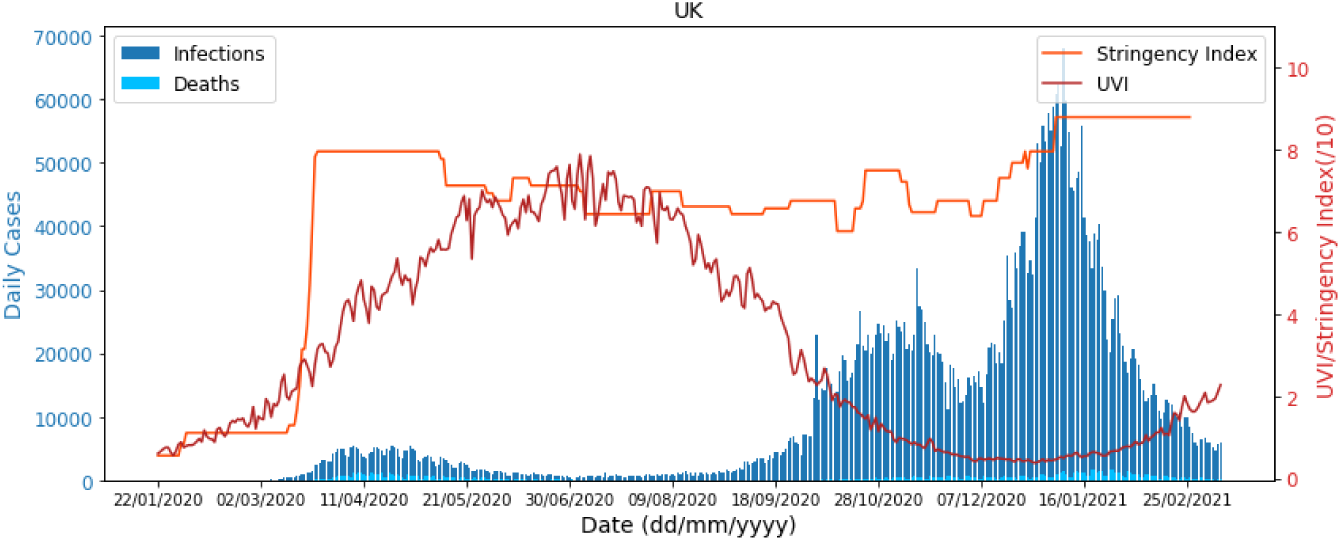
Daily infections (dark blue) and deaths (light blue) along with the stringency index (orange) and UVI (red) for the period 22 January 2020 up to 10 March 2021 for the UK. As expected, the UVI increased from January 2020 to July 2020 when it reached a peak and then dropped. From January to April 2020 while the UVI increased, the number of infections went up too. The UK government lockdown started on 23 March 2020 and this resulted in a decrease in infections due to social distancing. The lockdown was relaxed on 11 May 2020. However, increase in the UVI over the period 23 March to 1 July 2020 might have helped as well to decrease the number of infections. Over the period 2 July to October 2020 the increase in infections is strongly anti-correlated with UVI, as we quantify in Table 1. In November 2020 we see a decrease in the number of cases due to the national lockdown that was imposed, but then the cases increase again once the lockdown was lifted. During this period the UVI was low and therefore did not have an impact in reducing cases.

To quantify the correlation seen in Figure 1 we consider the correlation coefficient between X and Y, defined as

**Table 1:** The correlation coefficient *ρ* defined in Eq. 1 for different time intervals and time lags. Our logic in the selecting these time intervals is as follows: the first UK lockdown was between 23 March and 11 May 2020, and a second lockdown started a week after 28 October 2020. The minimum number of infections happened on approximately on 1 July. Formal bootstrap error bars on *ρ* are less than 0.05.

| Date Range | Corr. Coef. $\rho$ of $\log_{10}(UVI)$ & $\log_{10}(F)$ | | |
| --- | --- | --- | --- |
|  | No lag | 7 day lag | 14 day lag |
| 23 Mar 20 - 10 Mar 21 | −0.917 | −0.910 | −0.886 |
| 11 May 20 - 10 Mar 21 | −0.934 | −0.922 | −0.896 |
| 02 Jul 20 - 10 Mar 21 | −0.927 | −0.910 | −0.879 |
| 28 Oct 20 - 10 Mar 21 | −0.751 | −0.738 | −0.611 |

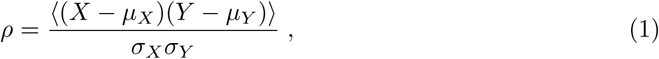

where *µ* represents the mean and *σ* the standard deviation. We applied this to *X* = log_10_(*UV I*) and *Y* = log_10_(*F*). We find the correlation coefficient averaged over the period 23 March 2020 (day that the first lockdown was imposed in the UK) to 10 March 2021 to be *ρ* = −0.917 suggesting an anti-correlation with a p-value of 1.80×10^−141^, implying that the null hypothesis of no correlation is strongly ruled out. Additionally, we calculate the correlation coefficient for the period 11 May 2020 (when initial relaxations of lockdown began) until the 10 March 2021 and find this to be *ρ* = −0.934 with a p-value of 2.24×10^−136^. ^1^ Over this period the stringency index was roughly constant to within 13%, so variations due to this factor were probably limited.

The correlation coefficient *ρ* for different date ranges and time lags of 7 and 14 days is shown in Table 1. Time lag makes small difference to *ρ*, probably as competing effects wash out a particular time lag. In Figure 2, the top panel shows the daily infections and UVI and on the bottom panel we plot the rolling correlation coefficient between log_10_(*UV I*) and log_10_(*F*) with a window size of 50 days. We observe a negative correlation between the infections and UVI from mid-April onwards.

**Figure 2:**
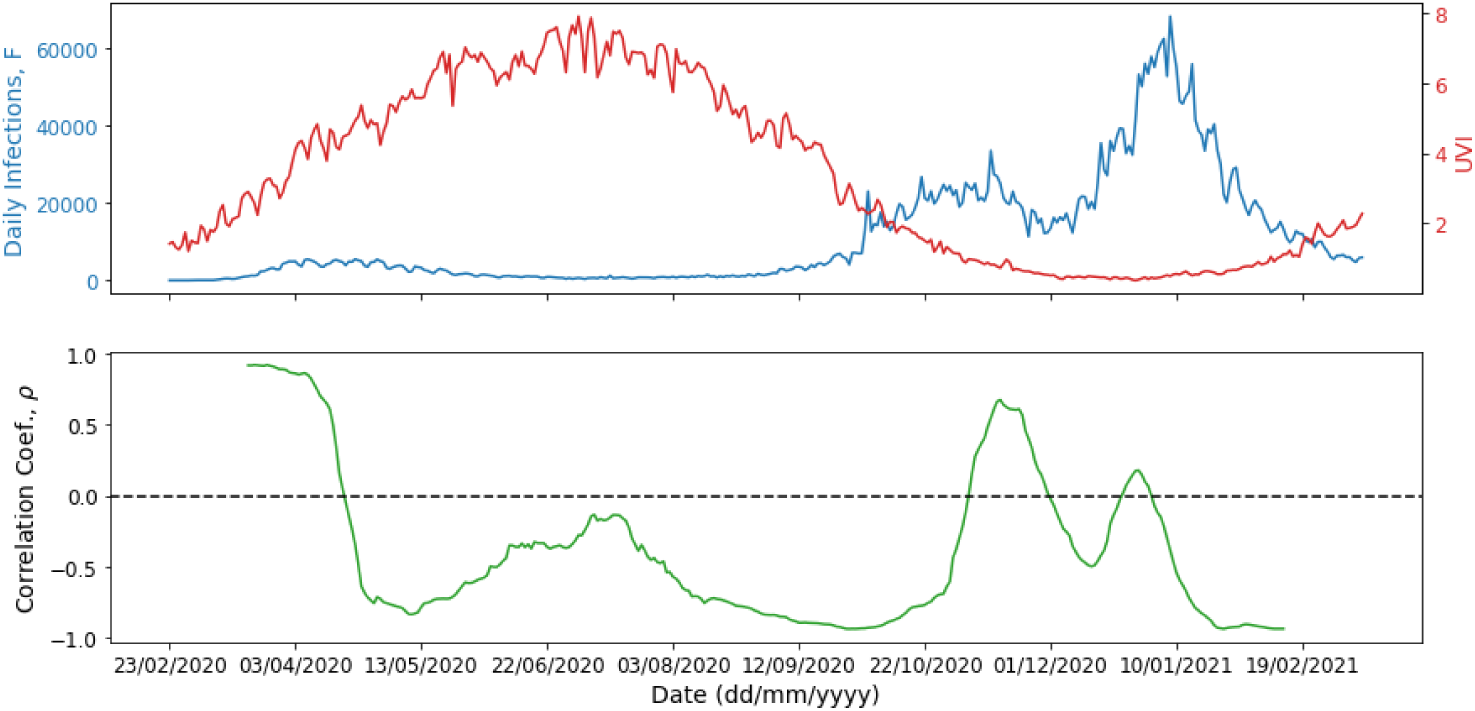
Top panel shows the daily infections (blue) and UVI (red) for the UK from the 23 February 2020 until the 10 March 2021. In the bottom panel we plot the rolling correlation coefficient of *log*_10_(F) and *log*_10_(UVI) (green) with a window size of 50 days.

Figure 3 shows a log-log scatter diagram of *F* vs. UVI, with colour coding for time intervals. The nearly linear correlation is remarkable. We fit the data to

**Figure 3:**
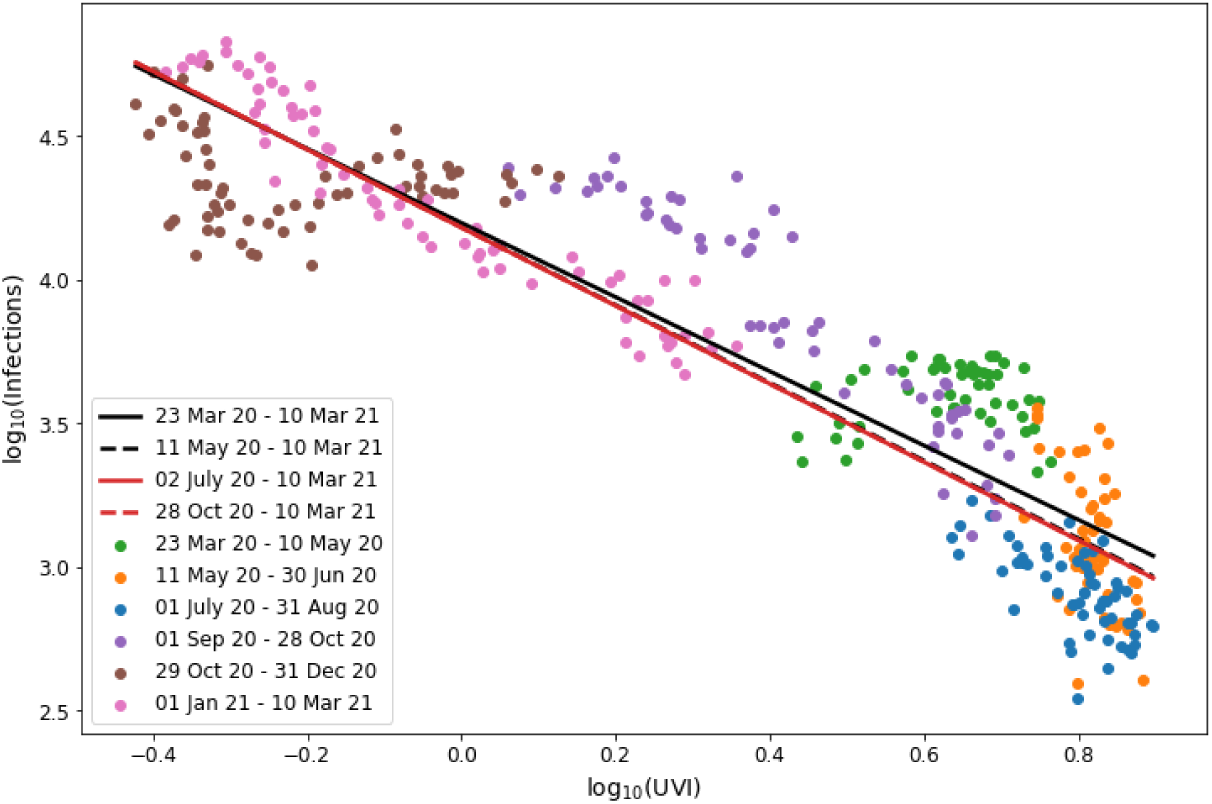
Log-log scatter diagram of F against UVI with colour-coded time intervals. The solid black line shows a fit by linear regression (Eq. 2) for the period 23 March 2020 to 10 March 2021 and the black dashed line for the period 11 May 2020 to 10 March 2021. The solid red line represents the period between 2 July 2020 to 10 March 2021 and the dashed red line the period 28 October 2020 to 10 March 2021.

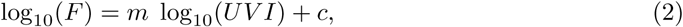

using standard least squares linear regression. For the period 23 March 2020 to 10 March 2021 we find find *m* = −1.292 *±* 0.029 and *c* = 4.199 *±* 0.017 where the 68% CL were derived using Bootstrapping [32]. The other three fits shown in Figure 3 are within 10% for the slope and 1% for the intercept.

## 4 Discussion

In conclusion, we have found an intriguing empirical anti-correlation between the daily UVI and COVID-19 infections in the UK, with a correlation coefficient of −0.934 between 11 May 2020 and 10 March 2021. We are in the process of extending our analysis for other countries and regions. For now we shall only mention the example of Chile, as a Southern Hemisphere case. Over the periods of rise (25 March to 6 June 2020) and fall (7 June to 28 October 2020) in infections we find an anti-correlation of *ρ* = −0.907 and *ρ* = −0.730 respectively, in accord with the anti-correlation we found for the UK. Another important test would be to contrast countries with the same environmental effects but different lockdown policies, for example Norway and Sweden.

We note that *a correlation* between two observables does not necessarily mean *causation*. If the UV light directly affects the level of infections it could be either by reducing the survival of the virus itself or by improving the immunity of people via the production of Vitamin-D and possibly a combination of the two. On the other hand, the UVI may indicate other causes. For example, low UVI may imply that people spend more time with other people indoors (e.g. at home or in shops) or suffer from other winter-time diseases, increasing the chance for infection. Other possible variables are population density, temperature, humidity, air pollution and other geographical parameters.

We emphasise that the infection by COVID-19 is a multi-parameter problem. There is indeed strong empirical evidence that keeping social distancing and wearing masks are the significant factors in reducing the transmission. It is also clear that the optimal solution to the infection would be via a vaccine. But in any case we advocate including the impact of the UV in the modelling of COVID-19 spread and in proposed medical protocols.

## Data Availability

We use publicly available data from JHU on COVID-19 and from TEMIS on UVI.

## Code availability

We plan to make the code available at a later stage.

## Acknowledgements

We thank R. Ellis, R. Lahav, C. La Porta, P. Lemos, D. Pillay, M. Treyer and S. Zapperi for helpful discussions. BH, OL and CN are supported by the STFC UCL Centre for Doctoral Training in Data Intensive Science (grant number ST/P006736/1) (ST/R000476/1).

## Author contributions

AB has proposed the idea of looking at correlations between the UVI and COVID-19 infections and has contributed to interpretation of results. CN and BH have performed the statistical analysis of this work. OL has proposed analysis tools and plots, has contributed to interpretation and has coordinated the team. Each of the four authors have written sections of the paper.

## Footnotes

1 We note that we identified two outliers in the number of daily infections which resulted from merging pillars of reported cases on the 1 and 2 July and hence were removed from the analysis.

